# Body composition and blood pressure relationship among western-Kenya school children and adolescents

**DOI:** 10.1101/2023.10.23.23297393

**Authors:** Karani Magutah, Grace Wambura Mbuthia, Rebecca M. Meiring

## Abstract

Kenyan studies relating body composition (BC) and blood pressure (BP) have focused on adults. Insight into this relationship among children and adolescents provides evidence for reduction of adulthood cardiovascular disease-risk development. We describe BC and BP correlation among Kenyan children and adolescents.

This cross-sectional study randomly sampled 389 school-going participants. Weight, height, BP, waist circumference (WC) and hip circumference were measured. Waist-to-hip ratio (WHR), waist-to-height ratio (WHtR) and body-mass-index (BMI) were calculated. Relationships between BP and WC, WHR, WHtR and BMI were determined.

Males and females were 14.7±2.9 and 14.4±2.5 years old, respectively. Secondary and primary schools’ males had SBP 119.1±10.3 mmHg and 107.4±13.7 mmHg; females 111.2±11.8 mmHg and 106.7±11.7 mmHg respectively. The DBP in secondary and primary schools’ males was 69.2±7.8 mmHg and 65.7±8.6mmHg, and, 69.6±7.9 mmHg and 68.1±7.4 mmHg for females. Higher BMI, WC, and WHtR correlated with higher SBP in both sexes (Pearson’s “R” +0.3 to +0.5; p<0.05). In females, DBP correlated with BMI, WC and WHtR, and only with WC for males. In all schools, only WC correlated with SBP in both sexes, BMI, WHR and WHtR correlating only among females. For males, BMI in primary and WHtR and WHR in secondary schools additionally correlated with SBP. For females, BMI and WHtR had correlations across school levels. For private and public schools, BMI, WC and WHtR had +0.2 to +0.6 correlations with SBP. The DBP correlated with WC and WHtR among private schools’ males, and, with WHtR and BMI in both school sets for females.

For both sexes, WC had strongest and WHR weakest correlation with SBP. Only in secondary school females did DBP correlate some BC variables. In both sexes, BMI, WC and WHtR all fairly correlated with SBP in private and public schools, WHR appearing least applicable in predicting Kenyan children and adolescents’ BP.

## Introduction

While much has been done on the relationship between body composition (BC) and blood pressure (BP), most research has focused on adults. Waist circumference (WC), body mass, waist-hip ratio (WHR), waist-height ratio (WHtR) and body mass index (BMI) are all either independently or in combination positively correlated with BP in adult males and females (1). What remains unclear is how various BC variables correlate with BP among the sexes and age groups. In Africa, childhood onset hypertension is currently on the rise (2). Among younger adults WC and WHR have more predictive power than BMI for occurrence of high BP, and, while WHR correlates more positively with Systolic BP (SBP), it is WC that correlates stronger with Diastolic BP (DBP) (3). Among children and adolescents, such relationships remain unresolved. We do not know as yet what BC variables confer the strongest association with higher BP for boys and girls, and, in Kenya, we additionally do not know exactly how the various BC components relate with BP for our children and adolescents.

The available studies focusing on BP and BC relationships in children and adolescents are all from outside Kenya and as far as we are aware, there is no literature available in this group in Kenya. Insight on the relationship between BP and the various BC variables in this group is needed, where available data are in adults and older youths aged 18-35 years, where BC factors like BMI ≥25 had 3.05 likelihood odds individuals were hypertensive (4). For those from primary school children only, up to 42% had elevated to stage 1 hypertension BPs and those overweight or obese had higher likelihood for elevated or hypertensive BP values (5). The current study provides the first attempt to describe the relationship between BC variables and BP in a western Kenya cohort of children and adolescents from both primary and secondary schools, extending the study age to 18 years.

Recent studies from elsewhere across the globe have shown mixed results in the relationship between various BC variables and BP in children and adolescents (6-9). In urban South Africa for instance, while weight, BMI and WC have positive correlations with both SBP and DBP, WHR showed no such correlation. In data disaggregated based on sex however, weight, BMI and WC correlate with SBP for both boys and girls but not in DBP among boys, and this is true for girls regards WC (8, 9). Other studies from both similar and different populations suggest that weight and BMI positively correlates with both SBP and DBP in boys and girls, and that high WHtR also compares with higher BP (7, 10). Studies have also shown that these factors though interrelated can independently cause higher BP in isolation even where the other variables of body composition are normal. Higher values of WC for instance correlates with higher BP even in individuals with normal BMI (11).

The aim of this study was to describe the correlation between BC variables and BP among children and adolescents from primary and secondary, and, also, private and public schools in Kenya.

## Methods

### Design and Setting

A cross-sectional observational study was conducted among residents of metropolis of Eldoret in North-West of Kenya.

### Study population

Children and adolescents aged 11-18 years and were residents of Eldoret town currently attending either primary or secondary schools under the Kenyan curriculum.

### Sample size and Sampling Procedure

A total of 389 participants were sampled from public and private schools, all drawn from the upper grades of primary school and the four secondary school levels.. This was done by listing all schools in each category (primary (private and public) and secondary (private and public) and writing their names on ballot papers, and from each, two were randomly picked to, yielding 8 schools. All agreed to participate and were included. Using the class registers from the selected schools, proportioned random sampling was employed based on the class size against the desired total sample, approximating a representation of 12 participants per grade in each school.

#### Eligibility

Children and adolescents aged between 11-18 years attending school within Eldoret were included. Children and adolescents with physical disabilities that could compromise engagement in PA and exercises, and those on medication that alters BP or BC were excluded.

#### Protocol Description and Data Collection

For this study, the first participant was recruited on 6th March 2023 and engagement with the last participant was on 28th April 2023. These children and adolescents were assented and their respective parents / guardians gave written-informed consent for before participation. A CAMRY Mechanical scale (BR9012, Shanghai, China) was used to measure body weight (kilograms) with participants wearing light clothing. Height was measured using a tape measure and taken while standing upright with eyes fixed directly ahead, and without wearing shoes. BP (mmHg) was measured after 5 minutes of rest in an upright-seated position. An Omron M2 Basic (HEM-7120-E) automatic BP monitor (Omron Healthcare Co. Ltd, Kyoto, Japan) was used to measure BP on the left upper arm. Measurements were taken twice, spaced two minutes apart, and the average of the two measurements was recorded. The WC was determined by “just snugly” passing a tape horizontally just above the iliac crest, with the lower part of tape barely touching the upper part of the crest and the hip circumference was taken as the measured distance around the widest part of the buttocks (12, 13). All tape-measure values were in centimeters. Using these measurements, BMI, WHtR and WHR were determined. The set cut-offs for WHtR were ≥0.463 for boys and ≥0.469 for girls (14-16), and ≥0.90 (male) and ≥0.85 (female) for WHR (13). We used a 95 percentile of adult values for BMI, applied amongst <13 year old, while those ≥13 years retained adult categorization (17, 18). Normal BP was taken as values <120/80 for ages ≥13 years (18).

### Data Management and Analysis

Analyses were performed using STATA v.13 at univariate level for BP, WC, WHR, WHtR and BMI data into means and standard deviations. For the relationship between the BC variables and both SBP and DBP, the bivariate Pearson’s correlations (“R”) were performed. Chi-squared tests were performed for BP comparisons between sexes. The P value for significance was set at ≤0.05.

### Ethical Consideration

The Moi Teaching and Referral Hospital / Moi University Institutional Research Ethics Committee (MTRH/MU IREC) approved this research (approval number FAN: 0004291) on 22^nd^ November 2022 and, further, the National Commission for Science, Technology and Innovation (*NACOSTI*) provided permit to carry out the study Ref number 839919 on 1^st^ March 2023. Written assent was provided by all children and adolescents and their parents/guardians gave informed-written consent for the participants to be enrolled.

## Results

For the current study, 37.4% of participants were male and 62.6% were female with mean±SD ages 14.7±2.9 and 14.4±2.5 years respectively. The proportions from secondary and primary schools were 46.9% (public, 33.8%; private 13.1%) and 53.1% (public, 22.6%; private, 30.5%) respectively. Participants were all from the 5^th^ grade (seven years in school) of primary schools up to 4^th^ form (Fourteen years in school) in the Kenyan education system. Of these participants, 7.6% (n=29) had hypertensive pressure ≥130 mmHg (≥124 mmHg for <13 year old) using SBP, 18.5% had prehypertensive SBP 120-129 mmHg (114-123 mmHg for <13 year old) and 73.9% had normal SBP. Using DBP, 7.3% (n=28) of participants were prehypertensive (80-89 mmHg (≥76-85 mmHg) for <13 years old)). Combining SBP and DBP, 8.1% (n=31) of participants (9.3% (n=13), males; 7.4%, females (n=18)) were prehypertensive-to-hypertensive and males had higher SBP compared to females (χ^2^ = 9.81; p = 0.007). Table 1 provides characteristics of the study participants per sex and school categories.

**Table 1.** Characteristics of Study Participants.

| Variable | Mean ± SD | | | |
| --- | --- | --- | --- | --- |
| Sex, School | Boys, Secondary Sch (n=62) | Boys, Primary Sch (n=84) | Girls, Secondary Sch (n=120) | Girls, Primary Sch (n=123) |
| Age (years) | 17.5±2.0 | 12.7±1.4 | 16.6±1.4 | 12.4±1.2 |
| Height (cm) | 170±8.2 | 154.8±11.2 | 160.5±8.2 | 154.5±9.8 |
| Weight (Kgs) | 60.7±9.3 | 42.1±10.6 | 57.6±11.6 | 44.4±10.7 |
| Waist (cm) | 79.0±7.5 | 68.5±7.8 | 75.6±10.3 | 66.9±8.6 |
| Hip (cm) | 91.6±7.3 | 79.7±8.5 | 97.0±9.1 | 84.6±9.3 |
| BMI (kg/m<sup>2</sup>) | 21.0±2.9 | 17.4±2.6 | 22.4±4.0 | 18.4±3.1 |
| SBP (mmHg) | 119.1±10.3 | 107.4±13.7 | 111.2±11.8 | 106.7±11.7 |
| DBP (mmHg) | 69.2±7.8 | 65.7±8.6 | 69.6±7.9 | 68.1±7.4 |
| WC (cm) | 79.0±7.5 | 68.5±7.8 | 75.6±10.3 | 66.9±8.6 |
| WHR | 0.86±0.07 | 0.86±0.06 | 0.78±0.07 | 0.79±0.06 |
| WHtR | 0.46±0.04 | 0.44±0.04 | 0.47±0.06 | 0.43±0.05 |

SD, standard deviation; cm, centimeters; Kgs, Kilograms; BMI, body mass index; kg/m<sup>2</sup>, kilogram per meter squared; SBP, systolic blood pressure; mmHg, millimeters of mercury; DBP, diastolic blood pressure; WC, waist circumference; WHR, waist hip ratio; WHtR, waist height ratio.

For both males and females, higher BMI, WC and WHtR all had significant positive relationships with SBP (all p <0.05). The WHR had significant positive relationship with SBP in females but not males. For DBP however, there were mixed results with BMI and WHtR having a significant positive correlation among females but not males, and, for both sexes, WC positively correlating with DBP while WHR had no correlation whatsoever. These correlations are presented in table 2.

**Table 2.** Pearson’s Correlations of various BC Variables to SBP and DBP.

| Variable (males (n=138) and females (n=241), respectively) | Correlation coefficient 'r' (p-value) |  |
| --- | --- | --- |
|  | SBP | DBP |
| BMI |  |  |
|  | 0.40 (<0.01) | 0.13 (0.13) |
|  | 0.32 (<0.01) | 0.30 (<0.01) |
| WC |  |  |
|  | 0.50 (<0.01) | 0.22 (0.01) |
|  | 0.43 (<0.01) | 0.27 (<0.01) |
| WHR |  |  |
|  | 0.10 (0.25) | 0.02 (0.79) |
|  | 0.31 (<0.01) | 0.12 (0.05) |
| WHtR |  |  |
|  | 0.27 (<0.01) | 0.16 (0.07) |
|  | 0.38 (<0.01) | 0.29 (<0.01) |

BMI, body mass index; SBP, systolic blood pressure; DBP, diastolic blood pressure; WC, waist circumference; WHR, waist hip ratio; WHtR, waist height ratio.

Correlating the various BC variable to both SBP and DBP based on sex and whether participants were in primary or secondary school levels had mixed results. Across both sexes and in all school levels, WC was the only one with a positive significant correlation with SBP. Additionally, BMI, WHR and WHtR all showed positive correlation to SBP among females in both primary and secondary schools. For males, other than WC, BMI in primary school boys and both WHR and WHtR in secondary school boys correlated with SBP. No BC variable correlated with DBP for males in all school levels. For girls, there was a correlation between DBP and all of BMI, WC and WHtR in secondary schools and BMI and WHtR for primary schools. This is shown in table 3.

**Table 3.**
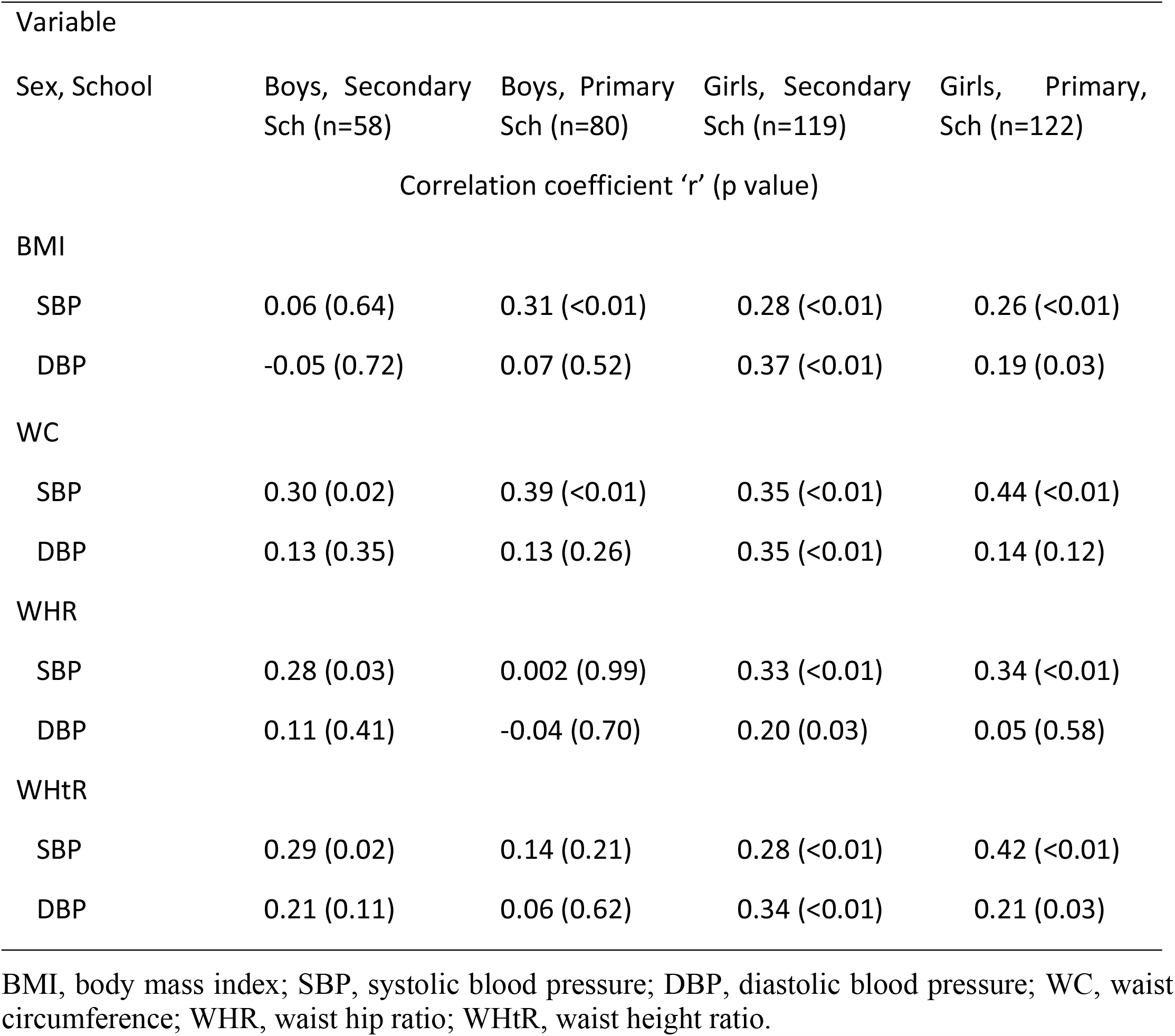
Pearson’s Correlations of various BC variables to SBP and DBP based on school level.

When analyses were based on whether participants were in private or public schools, BMI, WC and WHtR all positively correlated with the SBP. A significant correlation between WHR and SBP was only found in females from public schools. There were mixed results pertains DBP. Among males in private schools, WC and WHtR correlated with DBP, and there was no correlation for BC variables for those from public schools. The WC, WHtR and BMI correlated with DBP among females from both sets of schools. These correlations are presented in table 4.

**Table 4.** Pearson’s Correlations of various BC variables to SBP and DBP based on school type.

| Variable |  |  |  |  |
| --- | --- | --- | --- | --- |
| Sex, School | Boys, private sch (n=80) | Boys, Public sch (n=58) | Girls, private (n=86) | Girls, public (n=155) |
|  | Correlation coefficient 'r' (p value) |  |  |  |
| BMI |  |  |  |  |
| SBP | 0.41 (<0.01) | 0.39 (<0.01) | 0.37 (<0.01) | 0.33 (<0.01) |
| DBP | 0.16 (0.16) | 0.09 (0.49) | 0.29 (0.01) | 0.30 (<0.01) |
| WC |  |  |  |  |
| SBP | 0.41 (<0.01) | 0.60 (<0.01) | 0.30 (0.01) | 0.46 (<0.01) |
| DBP | 0.33 (<0.01) | 0.08 (0.53) | 0.25 (0.02) | 0.29 (<0.01) |
| WHR |  |  |  |  |
| SBP | -0.03 (0.76) | 0.23 (0.08) | 0.14 (0.19) | 0.34 (<0.01) |
| DBP | 0.06 (0.58) | -0.01 (0.92) | 0.23 (0.04) | 0.09 (0.25) |
| WHtR |  |  |  |  |
| SBP | 0.22 (0.04) | 0.35 (0.01) | 0.32 (<0.01) | 0.40 (<0.01) |
| DBP | 0.25 (0.02) | 0.01 (0.92) | 0.33 (<0.01) | 0.27 (<0.01) |

BMI, body mass index; SBP, systolic blood pressure; DBP, diastolic blood pressure; WC, waist circumference; WHR, waist hip ratio; WHtR, waist height ratio.

## Discussion

Although there were almost twice as many females as males in the current study, their ages were similar. The numbers drawn from secondary and primary schools, and, also, private and public schools were approximately equal. One in thirteen and 2 in 11 participants had hypertensive and prehypertensive SBPs respectively, and, using DBPs, 1 in 14 participants were prehypertensive. When the two BP categories were combined, 1 in 12 participants, a majority of whom were males were prehypertensive-to-hypertensive, with mean SBP in males higher than in females. This agrees with recent data from urban Kenya, although with participants drawn only from urban primary schools, that showed comparable proportions having elevated to stage 1 hypertension (5). Comparing males and females in either secondary or primary schools, males had higher SBPs while females had higher DBPs. This has been demonstrated before where mean SBP was significantly higher in boys than in girls with the mean DBP exactly opposite (10). The current study provides the first description BC variables have on SBP in school-going Kenyan children and adolescents at a time childhood onset hypertension is noted to be on the rise in Africa(2).

Using proposed correlation coefficients for interpreting medical data (19), WC, BMI and WHtR had fair (+0.3 to +0.5) correlations while WHR had no correlation with SBP among males. For females, WC, WHtR, WHR and BMI all showed fair (+0.3 to +0.4) positive correlation with SBP. In females, BMI, WC and WHtR all had fair (+0.3) positive correlation with DBP, and no correlation existed for WHR. Only WC had significant positive correlation with DBP among males, but this, too, was poor (+0.2). While some previous studies, although among older populations, found WC and WHR having higher correlation to high BP compared to BMI (3), the current study found BMI having a greater correlation compared to WHR in males but not in females. Like in Chaudhary et al (2019), WC had higher correlation to SBP compared to BMI for both sexes (R values 0.50 versus 0.40 for males and 0.43 versus 0.32 for females). Another area of divergence in the current study was that WHtR had higher correlation to higher SBP when compared to WHR in males and females. Like has been shown before, these BC variables correlated more with SBP than they did with DBP, and, where they did for DBP, it was only among females except for WC among males agreeing with Chaudhary et al (2019) when showing WC as the single most important variable for DBP correlation in older males and females (3). Elsewhere in urban South Africa, BMI and WC correlated with SBP for both boys and girls but not with DBP among boys (8, 9), largely agreeing with the current findings except for the diversion where the current study found BMI only and not together with WC as correlating with DBP among boys. Non-African studies, however, differ where weight, WHtR and BMI significantly correlated not only with SBP but also with DBP for both sexes (7, 10).

When data were disaggregated based on participants sex and whether they were in primary or secondary schools, there were mixed results. There was fair positive correlation between WHtR and WC on one side and SBP on the other for males in secondary schools while for those in primary schools, this fair correlation was instead seen in BMI and WC. For the females, all of BMI, WC, WHR and WHtR significantly correlated fairly with SBP in both primary and secondary schools alike. It was evident that across both sexes and in all school levels, WC was the only BC variable providing a positive significant correlation with SBP, which could be could explained from recent findings that WC has a positive association to arterial stiffness and that it affects vascular health directly (20, 21). The finding on WC could be utilized for both males and females in primary and secondary schools to predict whether they are likely to have a higher SBP. Among females in either primary or secondary schools, that BMI, WC, WHR and WHtR all showed positive significant correlation to SBP shows their predictive value in SBP level estimation in such groups, an application absent in males. Higher BMI among primary school children has been demonstrated to contribute to elevated-to-stage-1 hypertension in nearly half of previously studied cohorts (5). For males, it appeared only WC would be applicable for boys in both school category, with BMI only applicable in primary schools and both WHR and WHtR applicable among secondary school boys, complicating wholesome application. For both school categories, no BC variable correlated with DBP among males while among girls in secondary schools, the correlation between BMI, WC and WHtR on one side and DBP on the other was fair while that between WHR and DBP was poor though statistically significant. Among primary school girls, BMI and WHtR had poor but significant correlations with DBP, while no correlations were observed between DBP and either WC or WHR. Similar disparities have previously been demonstrated between sexes where WC for instance has been demonstrated as a more powerful predictor of higher BP in males and not females (22, 23).

Considering whether participants in the current study were from private or public schools, BMI, WC and WHtR all positively correlated (fair; +0.3 to +0.6) with the SBP for both boys and girls from both sets of schools. Higher BMI has been shown to correlate with higher BP in public schools’ adolescents in Cameroon (24), but there is paucity of comparable data for private schools from the African region. The WHR had no correlation with SBP in boys and only in females from public schools did it manifest a fair and significant correlation. Such inconsistencies involving WHR correlation with SBP have been described in other studies although involving private school children from outside Africa (25), only. While all three of BMI, WC and WHtR appear applicable in predicting how high SBP in such a cohort as the current might be regardless of what type of school participants attend, it looks feasible to say that WHR would only be applicable in females attending public schools, weakening its applicability over the rest. On the other hand, using BC to predict DBP among children and adolescents in school is not as straightforward as it appears for SBP. While for instance in private schools the WC and WHtR correlated with DBP among males in a fair range, no BC measurement correlated with DBP in boys from public schools. The WHtR, WC and BMI significantly correlated with DBP among females from private and public schools alike. Here, too, the WHR appeared unrealistic for use in predicting DBP in males from both school types and had mixed results among females, being appearing applicable in private but not public schools.

### Limitations

Two research assistants performed the BC measurements and this may have contributed some individual differences that affected the results that we could not control for, despite having trained the assistants together. Additionally, the BP measurements were done in a single session and using different (though calibrated) BP machines, and this may have affected conclusions on classification based on the once-off values.

## Conclusion

Among children and adolescents of both sexes and school levels, WC correlated strongest with SBP with WHR weakest in males and females, underlining its applicability in predicting SBP in Kenyan children and adolescents. For DBP, only in females especially in secondary schools were the BC variables correlatable, limiting their application in DBP prediction among males and primary school females. The BMI, WC and WHtR all fairly-to-moderately correlated with the SBP for both sexes from private and public schools and appear applicable for predicting SBP in such cohorts, while WHR may only be applicable in females attending public schools. Use of BC to predict DBP had mixed results for private versus public schools, but WHR was clearly unrealistic for use in predicting DBP in both sexes and in both school categories except, weakly, for girls in private and secondary.

## Data Availability

All relevant data are within the manuscript and its Supporting Information files.

## Acknowledgement

KM was supported by the Consortium for Advanced Research Training in Africa (CARTA). CARTA is jointly led by the African Population and Health Research Center and the University of the Witwatersrand and funded by the Carnegie Corporation of New York (Grant No. G-19-57145), Sida (Grant No:54100113), Uppsala Monitoring Center, Norwegian Agency for Development Cooperation (Norad), and by the Wellcome Trust [reference no. 107768/Z/15/Z] and the UK Foreign, Commonwealth & Development Office, with support from the Developing Excellence in Leadership, Training and Science in Africa (DELTAS Africa) programme. The statements made and views expressed are solely the responsibility of the Fellow.

